# Characterizing the skeletal muscle immune microenvironment for sarcopenia using transcriptome analysis and histological validation

**DOI:** 10.1101/2024.02.23.24303270

**Authors:** Linhui Shen, Yuan Zong, Jiawen Zhao, Yi Yang, Lei Li, Ning Li, Yiming Gao, Xianfei Xie, Qiyuan Bao, Liting Jiang, Weiguo Hu

## Abstract

Sarcopenia is a condition characterized by the age-related loss of skeletal muscle mass and function. The pathogenesis of the disease is influenced by chronic low-grade inflammation. However, the specific changes in the immune landscape changes of sarcopenic muscle are not yet fully understood. To gain insights into the immune cell composition and interactions, we combined single-nuclei RNA sequencing data, bulk RNA sequencing datasets, and comprehensive bioinformatic analyses on skeletal muscle samples from young, aged, and sarcopenic individuals. Histological staining was then performed on skeletal muscles to validate the distribution of immune cells in clinical samples. Overall, we analyzed the transcriptomes of 101,862 single nuclei, revealing a total of 10 major cell types and 6 subclusters of immune cell types within the human skeletal muscle tissues. Among the immune cells, macrophages constituted the largest immune fraction. A specific marker gene LYVE1 for skeletal muscle macrophages was further identified. Cellular subclasses included four distinct groups of resident macrophages, which play a different role in physiological or non-physiological conditions. Using bulk RNA sequencing data, we identified strong enrichment for a macrophage-rich inflammation in sarcopenia. Our findings demonstrate age-related changes in the composition and cross-talk of immune cells, which contribute to chronic inflammation. Furthermore, macrophages emerge as a potential therapeutic target, thus advancing our understanding of the pathogenesis of sarcopenia.

## Introduction

Sarcopenia is a systemic skeletal muscle disease that occurs with aging, characterized by the progressive loss of muscle mass and function(Cruz-Jentoft & Sayer, 2019). It is commonly associated with weakness, immobility, and premature death(Cruz-Jentoft, 2016). Similar to other human tissues, skeletal muscle degeneration also happens with age(Cruz-Jentoft & Sayer, 2019). The incidence of sarcopenia increases as individuals get older, with an average onset age of over 70 years old(Choe et al., 2022; Huang et al., 2021; Yu et al., 2014). This has made sarcopenia a significant public health concern in aging societies(Chen et al., 2020), as it is linked to numerous adverse outcomes such as falls, fractures, and mortality in the elderly. Histopathologically, sarcopenia is characterized by a reduction in the number and size of muscle fibers, especially Type-II muscle fibers, as well as fatty infiltration of skeletal muscle(Cruz-Jentoft & Sayer, 2019; Damluji et al., 2023). Muscle anabolic resistance, mitochondrial dysfunction, inflammation, and degenerative changes in the nervous system play important roles in the development of the disease(Damluji et al., 2023). However, the precise mechanism underlying the development of sarcopenia is not yet fully understood.

In recent years, there has been a growing interest in studying the relationship between sarcopenia, immunity, and inflammation. As individuals age, they often experience a chronic low-grade pro-inflammatory state characterized by and increase in pro-inflammatory cytokines and a decrease in immune cell function(Wilson, Jackson, Sapey, & Lord, 2017). Frail elderly individuals have higher levels of tumor necrosis factor-α (TNF-α) compared to healthy young men and women, and interleukin-6 (IL-6) is significantly associated with sarcopenia(Greiwe, Cheng, Rubin, Yarasheski, & Semenkovich, 2001; Langkilde et al., 2015). The elevated expression of these cytokines can contribute to muscle atrophy(Crossland, Constantin-Teodosiu, Gardiner, Constantin, & Greenhaff, 2008; Yakabe et al., 2018). Additionally, B cells may play an important role in the regulation of sarcopenia through transcriptional mechanisms(Zhang et al., 2023). Moreover, there is evidence of a correlation between sarcopenia and autoimmune diseases. For instance, rheumatoid arthritis (RA) patients are at an increased risk for sarcopenia(Bennett, Pratt, Dodds, Sayer, & Isaacs, 2023).Other autoimmune diseases, such as multiple sclerosis, celiac disease, type 1 diabetes, psoriasis, and ulcerative colitis have also been linked to sarcopenia(Jones et al., 2020). Therefore, further investigation into the interplay between sarcopenia, immunity, and inflammation is warranted.

Chronic inflammation in sarcopenia is a complex process involving various specialized immune cell types. The interactions between immune cells and native cells in skeletal muscle tissue are crucial for both the progression of inflammation and the repair of damage. In patients with sarcopenia, T helper 17 (Th17) cells are down-regulated, while NK cells are up-regulated. Furthermore, certain key genes regulate the progression of sarcopenia by influencing the immune microenvironment(Shen et al., 2023). When muscle is damaged, resident neutrophils and macrophages become activated, and the cytokines and growth factors released by activated T cells play a pivotal role in the proliferation and migration of muscle satellite cells(Saini, McPhee, Al-Dabbagh, Stewart, & Al-Shanti, 2016). However, the number of these cells decreases in older adults, potentially leading to inadequate muscle repair. Despite several studies focusing on specific cell populations in sarcopenia, comprehensive analyses are still lacking.

Single-cell RNA sequencing (scRNA-seq) is a powerful technology that enables precise the measurement of gene expression at a cellular level. It provides valuable insights into the status of different cells in the body’s microenvironment, which cannot be achieved by traditional transcriptomic technologies. Due to challenges with skeletal muscle dissociation and cellular filtration, there are not many studies on the single-cell atlas of skeletal muscle system to date. Some related analyses have recently been published(Cho, Schmitt, Dasgupta, Ducharme, & Doles, 2020; Coulis et al., 2023; Giordani et al., 2019; Xu et al., 2021), but they still have a limited scope. In the present study, our aim was to explore the complete immune microenvironmental landscape of skeletal muscle at different ages based on single-nuclei RNA sequencing data and bulk RNA sequencing datasets. We also examined the gene expression signature of sarcopenia and the important role of immune cells in its progression. The identification of key genes and cells may provide a potential therapeutic target for intervening in the progression of sarcopenia.

## Results

### Cell type composition of skeletal muscle tissue from young and aged people

The single-cell dataset from GSE167186 included skeletal muscle tissue samples from 6 young and 11 old individuals. Through snRNA-seq analysis, we were able to identify the age-related changes in the cellular composition of skeletal muscle tissue. After performing quality control, we retained 101,862 nuclei for further analysis. Unsupervised classification partitioned the 101,862 nuclei into 16 clusters (**Figure 1A**). Based on the canonical gene markers, we manually assigned these clusters to 9 cell types: Type IIa, Type I, LUM+ Fibro-Adipogenic Progenitor (FAP) Cells, Endothelial Cells, Type II_2, ls, Pericytes, Immune cells and FBN1+ FAP Cells (**Figures 1B and D**). The study identified a cluster that exhibited characteristics of both Type I and Type II, referred to as mixed Type I/II. Differences in the numbers and distribution of different cell clusters in aged and young muscle, as well as the expression level and distribution of gene markers in immune cells, type I and type II muscle fiber were illustrated (**Figure 1C**). The volcano diagram in **Figure 1E** displayed the top differentially expressed genes (DEGs) in each cell type, revealing extensive transcriptomic alterations in cell function and metabolism in the aging muscle. The heatmap presented the top-enriched genes in each cluster (**Figure 1F**). Type I, and type II muscle cells were observed to be the main components in skeletal muscle tissue, although their composition varied between young and aged individuals (**Figure 1G**). Notably, the proportion of type IIa muscle cells showed a general downward trend in the aged group, and there was a significant decrease in endothelial cells, satellite cells, and pericytes compared to the young group. Although not statistically significant, the number of immune cells in the aged group exhibited an increasing trend (**Figure 1H**). The decreased number and fibrotic transformation of satellite cells in aging muscle might contribute to a decreased muscle regeneration capacity(Y. Wang et al., 2019). Previous studies have demonstrated a decrease in angiogenesis and blood flow in aged muscle tissue, which is associated with the development of sarcopenia(Das et al., 2018). Further investigations are needed to understand the changes in immune cell subsets and metabolism in the muscle aging microenvironment, considering the role of chronic inflammation in sarcopenia development.

**Figure 1.**
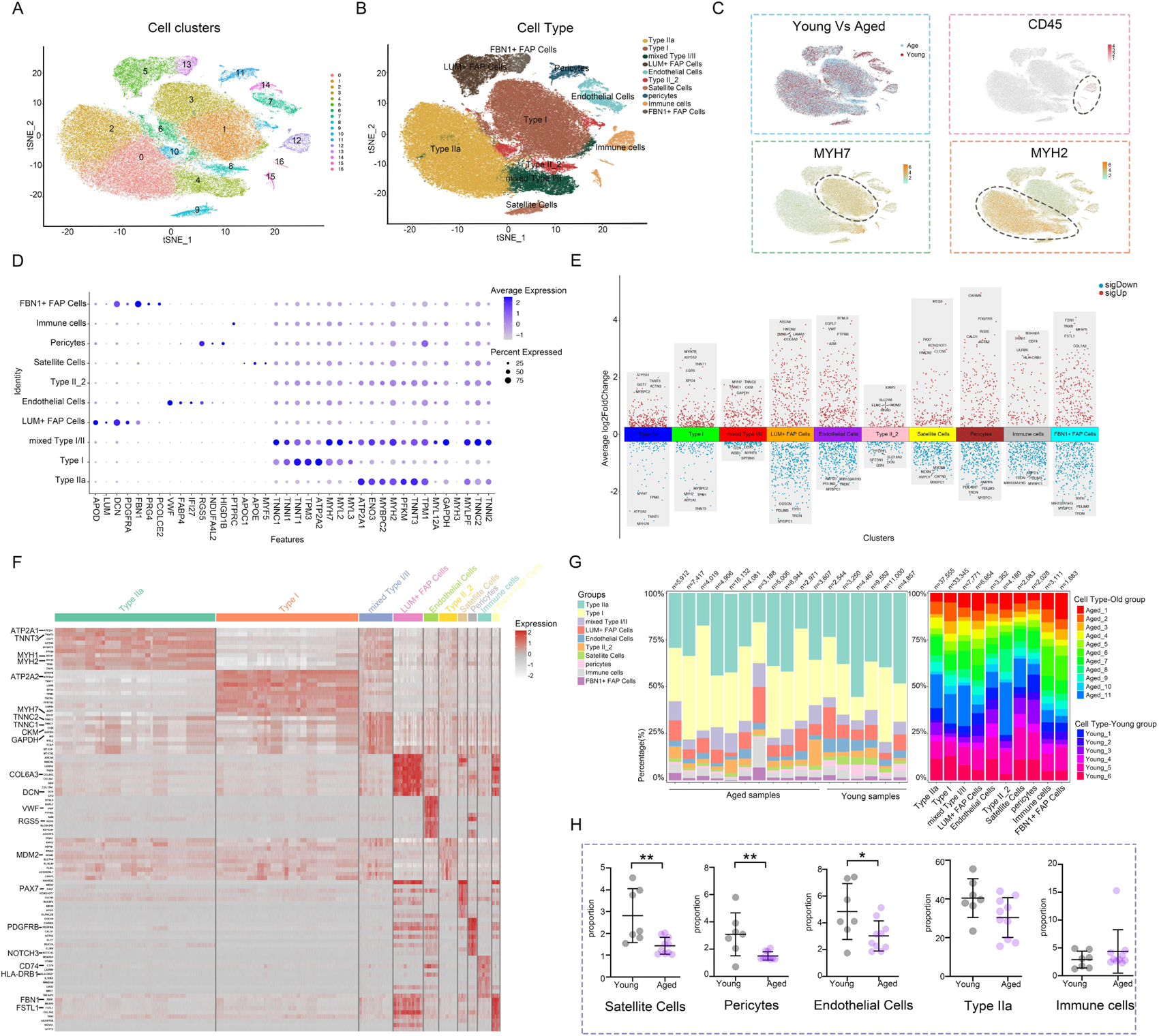
Analysis of the cell composition of young and aged skeletal muscle based on snRNA-seq data. **(A)** t-distributed stochastic neighbour embedding (t-SNE) plot of joint analysis of cell composition in young and aged skeletal muscle tissue. 16 unsupervised clusters were generated from 101,862 cell nuclei after quality control (distinguished by different colours and numbers). **(B)** Following dimensionality reduction, 10 clusters were manually annotated based on marker gene expression and visualized with t-SNE. **(C)** Feature plots (t-SNE) showing expressions of major genes that identify immune cell subpopulations. **(D)** Dot plots representing the percentage and average expression of selected marker genes for each cluster. **(E)** Differentially expressed genes (DEGs) in each cell type in aged versus young skeletal muscle. **(F)** Heat map depicting the expression of the top 10 upregulated genes identified in each cell cluster. Each row represents a single cell, and each column represents a single gene. **(G)** Bar plot depicting the proportion of each cell subset in each sample and the proportion of each sample in each cell subset. **(H)** Cell proportion analysis of satellite cells, pericytes, endothelial cells, Type IIa and immune cells between the young and aged (*p<0.05; **p< 0.01).

### Immune cell composition in skeletal muscle tissue

In order to explore the role of immune cells in the aging muscle microenvironment, we further grouped immune cells into 6 clusters: M2 macrophage, T cell, NK cell, M1 macrophage, mast cell and dendritic cell (DC) (**Figure 2A**). The expression profiles of marker genes for different cell populations were depicted in a violin plot (**Figure 2B**). Subsequently, we evaluated the interactions among immune cells. In the skeletal muscle microenvironment, a significant positive correlation was observed between DCs and M1 macrophages, NK cells and T cells, while a negative correlation was observed between DCs and mast cells (**Figure 2C**), which aligned with the proximity depicted in the UMAP. Furthermore, various functional and metabolic pathways were found to be enriched in different subtypes of immune cells. GO and KEGG pathway analysis revealed enrichment for terms such as collagen fibril organization, DC differentiation, JAK-STAT signaling pathway, endocytosis and TGF-β signaling pathway in M1-like macrophages. Similarly, T cells showed enrichment in terms related to skeletal muscle cell differentiation, fatty acid metabolism, cysteine and methionine metabolism. The NK cell population exhibited high enrichment in cytolysis and ribosome, while DCs showed enrichment in sphingolipid metabolism and natural killer cell mediated cytotoxicity (**Figures 2D and E**). The skeletal muscle immune microenvironment was a complex milieu consisting of several innate and adaptive immune cells, with innate immune cells being the majority. Among the innate immune cells, M1 and M2 macrophages were the most abundant (**Figure 2F**). Furthermore, the expression levels of some cell-specific marker genes involved in the immune response and aging process showed differences between the young and aged groups (**Figure 2G**). For instance, the aged group exhibited increased expression of CD226 and ADAM58 in DCs, CDKN1A in M1 macrophages, and NCR1, CD226 and KLRD1 in NK cells. Histological staining revealed an augmentation in immune cells within the aged samples (**Figure 2H**). HE staining indicated that the interstitial tissue of aged muscles exhibited a loose and fragmented structure, accompanied by a higher distribution of lymphocytes, while IHC staining confirmed the higher presence of T (CD3^+^), NK (CD56^+^), and hematopoietic stem and progenitor (CD117^+^) cells in the aged skeletal muscle samples. Given the critical position of the skeletal muscle immune microenvironment, we next sought to investigate the differences in macrophage expression and functions between the young and aged groups.

**Figure 2.**
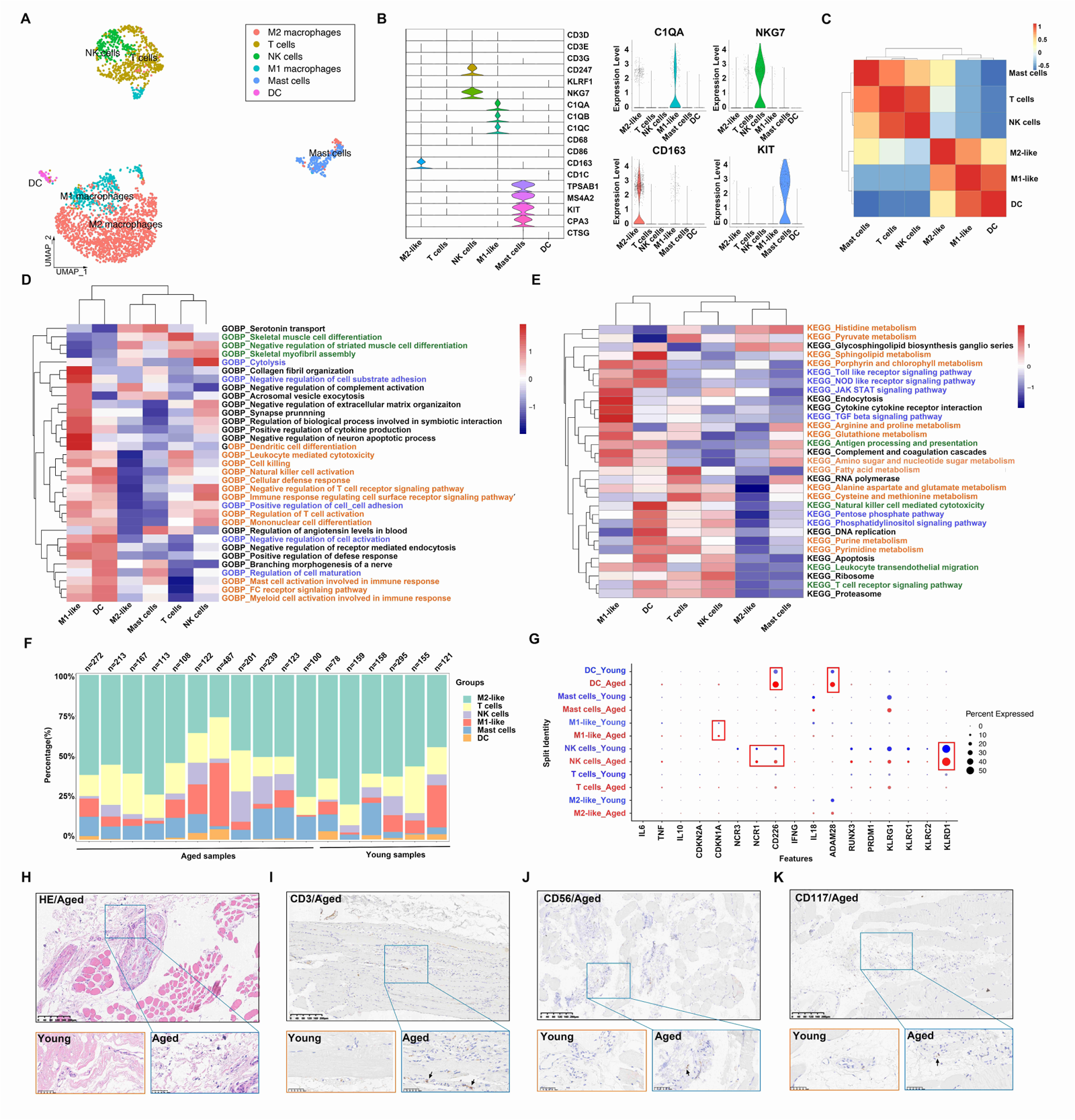
Analysis of the immune cell subpopulations of young and aged skeletal muscle based on snRNA-seq data. **(A)** Uniform manifold approximation and projection (UMAP) plot of 6 immune cell types for joint analysis in young and aged skeletal muscle tissue. **(B)** Violin plots of expression values for cell type-specific marker genes. **(C)** Heatmap of Spearman’s correlations among immune cell subtypes. Correlation coefficient values are colour-coded from blue (-0.5) to red (1.0) using the colour bar on the right. **(D-E)** Comparison of pathway enrichment across immune subpopulations. Heatmap of enrichment scores of selected Gene Ontology (GO) **(D)** and kyoto encyclopedia of genes and genomes (KEGG) **(E)** pathways in immune cell clusters following gene set variation analysis (GSVA). **(F)** Bar plot depicting the proportion of each immune cell subset in each sample. **(G)** Dot plots demonstrating the expression differences of inflammation-associated genes in each cluster in the young group versus aged individuals. **(H)** Representative images of hematoxylin and eosin (H&E) staining in young and aged skeletal muscle specimens. **(I-K)** Representative images of immunohistochemical (IHC) staining for CD3 **(I)**, CD56 **(J)** and CD117 **(K)** (scale bar = 100 μm).

### Macrophage subpopulations in skeletal muscle tissue

Macrophages were classified into four subgroups based on the expression levels of the selected marker genes LYVE1, HLA-DRB1, HLA-DRA and TSPO. These subgroups were LYVE1^lo^MHC-II^hi^, LYVE1^hi^MHC-II^lo^, LYVE1^lo^MHC-II^lo^ and LYVE1l^lo^MHC-II^lo^TSPO^+(Dick^ ^et^ ^al., 2019; Jin^ ^et^ ^al., 2022)^ (**Figures 3A and B**). A violin plot was used to visualize the gene signatures expressed by these subpopulations (**Figure 3C**). Subsequently, GO and KEGG analyses were conducted to investigate the functions of the four subpopulations. The results revealed that LYVE1^lo^MHC-II^hi^ was highly enriched in pathways related to phagosome, antigen processing and presentation, and macrophage, neutrophil, and B cell activation in immune response pathways. This suggests that LYVE1^lo^MHC-II^hi^ macrophages play an active role in the immune response. In contrast, LYVE1^hi^MHC-II^lo^ showed significant enrichment in pathways related to endocytosis, amino acid biosynthetic process and cellular response to glucose starvation. LYVE1^lo^MHC-II^lo^ exhibited upregulation in pathways associated with cardiac muscle contraction, skeletal myofibril assembly, acetyl CoA metabolic process and fatty acid metabolic process pathways. On the other hand, LYVE1l^lo^MHC-II^lo^TSPO^+^ displayed upregulation in pathways related to focal adhesion, protein digestion and absorption, ECM-receptor interaction, PI3K-Akt signaling pathway, collagen fibril organization, and response to fibroblast growth factor, indicating aclose connection with muscle fibrosis (**Figures 3D and E**). The significantly altered genes within each cluster were included as features for further functional enrichment analysis. The results revealed that innate immune response was associated with LYVE1^lo^MHC-II^hi^, while muscle structure development was associated with LYVE1^lo^MHC-II^lo^, and extracellular matrix organization was associated with LYVE1l^lo^MHC-II^lo^TSPO^+^ (**Figure 3F**). **Furthermore,** LYVE1^hi^MHC-II^lo^ macrophages were found to be the predominant component of macrophages in the skeletal muscle microenvironment (**Figure 3G**). Meanwhile, the LYVE1^hi^MHC-II^lo^ subpopulation exhibits an increasing trend in the macrophage composition of aging muscles (**Figure 3H**).

**Figure 3.**
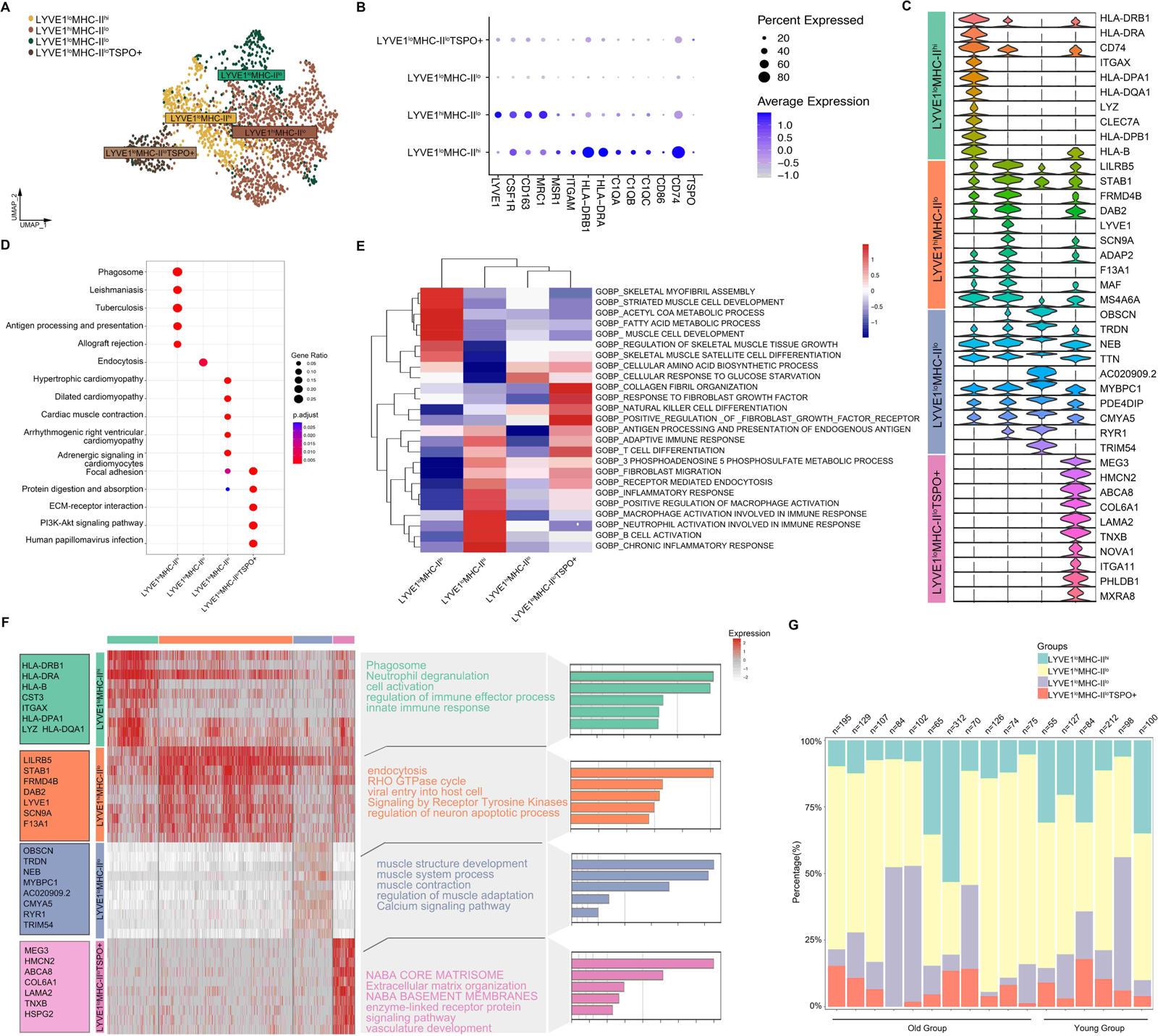
Deep profiling of the macrophage subpopulations in skeletal muscle microenvironment based on snRNA-seq data. **(A)** UMAP plot of 4 macrophage subtypes in skeletal muscle tissue. **(B)** Dot plots representing the expression levels of selected marker genes for each macrophage cluster. **(C)** Violin plots of expression values for selected marker genes for each cluster. **(D)** KEGG enrichment analysis revealed biological processes associated with each subset. **(E)** Heatmap of enrichment scores of selected GO pathways in macrophage clusters following GSVA. **(F)** Heat map depicting the expression of the top upregulated genes identified in each macrophage cluster. Each row represents a single cell, and each column represents a single gene. The left side displays the specific gene names, and the right side lists the pathways associated with these genes (inferred utilizing Metascape). **(G)** Bar plot depicting the proportion of each macrophage subset in each sample.

### Cell-cell communication in muscle immune microenvironment

The numbers and strength of interactions between major cell types and immune cells were found to be lower in the aged group, indicating changes in signaling pathways in the microenvironment of aged skeletal muscle (**Figure 4A-B**). Specifically, the aged group showed a decrease in the interactions between endothelial cells and Type II muscle fibers, while there was a significant increase in the f signaling patterns received by FBN1+ FAP Cells. Additionally, a unique signaling pathway was discovered, where pericytes sent signals to and received signals from immune cells (**Figures 4C and E**). When focusing on immune cells, overall interactions between immune cell types decreased in the aged group, but a new strong communication from M1 macrophages to T cells emerged, and communication from DCs to M2 macrophages also emerged (**Figures 4D and F**). Next, we performed IF staining on the walls of small blood vessels using α-SMA. Our findings revealed a significant increase in the number of CD45-labeled macrophages in the aged group compared to the young group. Furthermore, we observed a close association between the distribution of these macrophages and small blood vessels (**Figures 4G**). In general, the interaction between cell populations in aged muscle exhibited a decrease compared to the young group, suggesting that the aged microenvironment disrupts normal intercellular communication.

**Figure 4.**
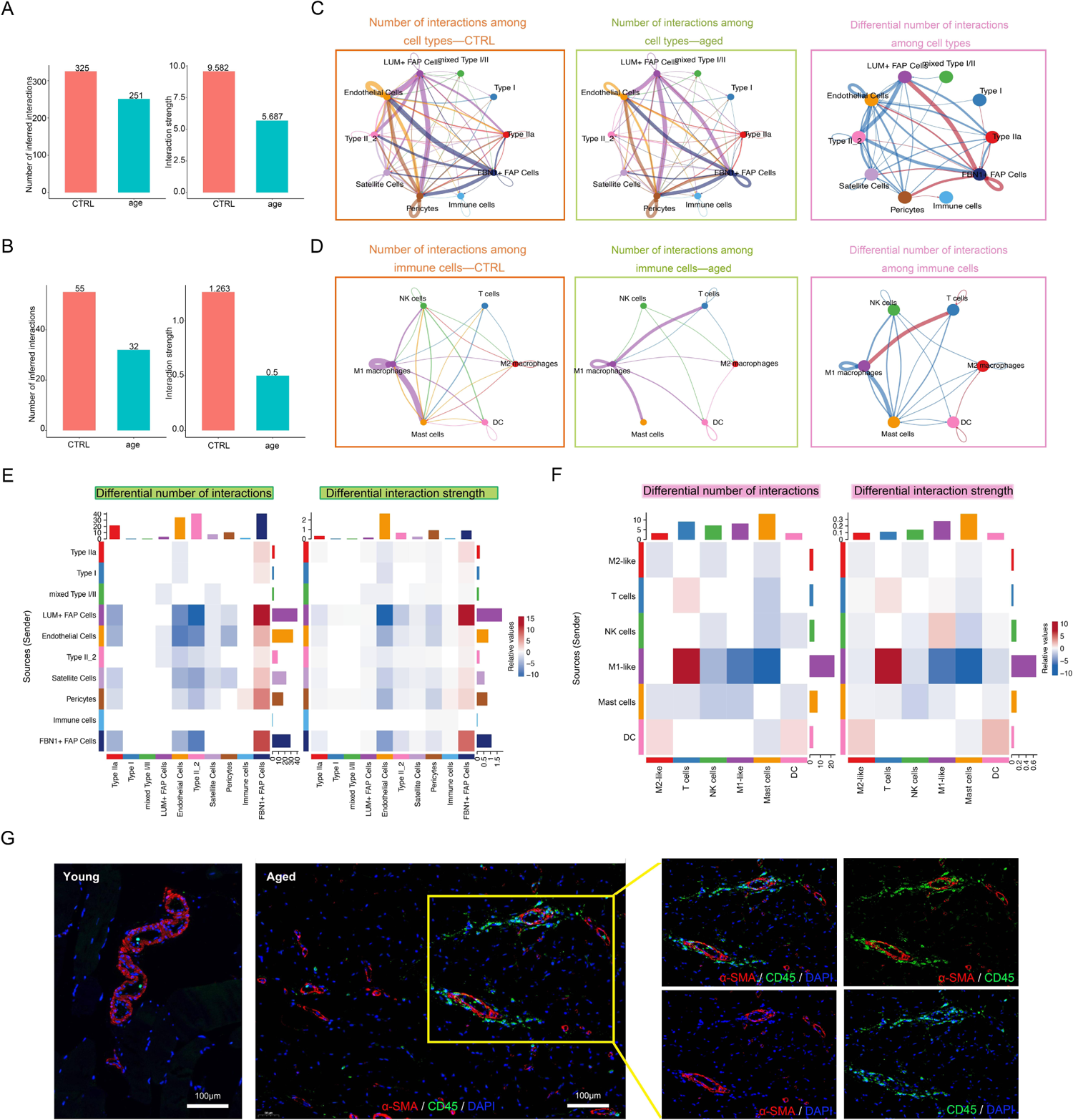
Cell-cell communication in muscle immune microenvironment. **(A-B)** Intercellular ligand–receptor prediction among cell types and immune cells revealed by CellChat. Bar plot showing the number and strength of intercellular interactions among cell types **(A)** and immune cells **(B)**. **(C-D)** Circle plots demonstrating the overview of cell–cell interactions among cell types **(C)** and immune cells **(D)**. Arrow and edge colour indicate direction and sender. Edge thickness reflects the number and the strength of interaction between populations. Differential number of interactions in the cell–cell communication network with red or blue coloured edges representing increased or decreased signaling in the aged group compared to the young group. Line thickness and darkness indicate the relative enrichment value. **(E-F)** Heatmaps of differential number and strength of intercellular interactions among cell types **(E)** and immune cells **(F)**. **(G)** Representative images of co-IF analysis of immune cells (CD45, green), blood vessels (α-SMA, red) and nuclei (DAPI, blue) of skeletal muscle tissues from the young and aged (scale bar = 100 μm).

### The enrichment of inflammatory signaling pathways in the sarcopenic tissue microenvironment

In this study, we utilized bulk RNA-seq to investigate the functional and metabolic changes between individuals with sarcopenia and those without. Several pathways were significantly enriched in patients with sarcopenia, such as linoleic acid metabolism, TGF-β signaling and cell cycle pathways, indicating metabolic and immune changes. On the other hand, compared to the young group, several pathways were downregulated in the sarcopenia group, including fructose and mannose metabolism, galactose metabolism and oxidative phosphorylation pathways (**Figure 5A**). To further understand the disease process of sarcopenia, we performed GSEA on RNA-seq data and identified additional biological pathways that could contribute to sarcopenia. The GSEA results confirmed enrichment of KEGG genes in cell cycle, JAK-STAT signaling pathway, ERBB signaling pathway and alpha-linolenic acid metabolism in the sarcopenia group (**Figure 5B**). We also observed enrichment of WIKI-pathway genes in cell cycle, as well as other pathways such as DNA IRdamage and cellular response via ATR, oxidative stress response and IL-4 signaling pathway (**Figure 5C**). Moreover, we investigated the gene expression of inflammation-related pathways and pathways related to muscle function. The results revealed significantly higher expression of genes associated with ERBB, IL-2, IL-4, oxidative stress response and JAK-STAT signaling pathway in the sarcopenia group (**Figures 5D and E**). Interestingly, these genes were down-regulated in the aged group and up-regulated in the sarcopenic group compared to young individuals. Furthermore, the correlation analysis revealed that most genes had a significant negative correlation with FAP cells and macrophages, while satellite cells, Type IIa, and Type I showed an overall opposite trend (**Figure 5F**). In summary, the findings suggest that sarcopenia is strongly associated with increased inflammation and impaired muscle function, which is linked to FAP cells and macrophages.

**Figure 5.**
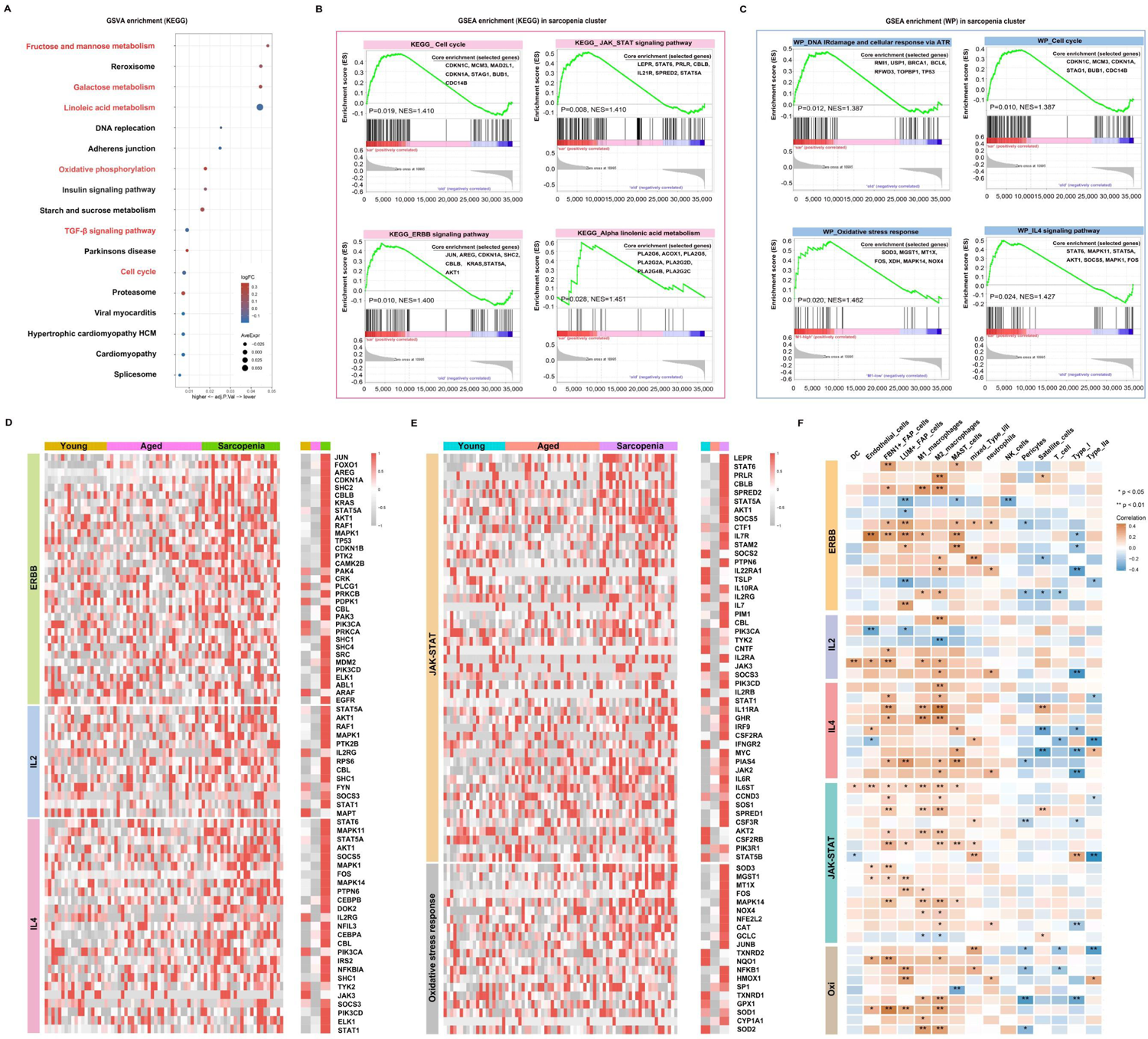
inflammation is a vital pathophysiological mechanism for sarcopenia. **(A)** Visualization of GSVA for pathways enriched in samples from patients with sarcopenia as compared to samples without sarcopenia using a bubble plot with the color coding indicating the −log10 (p adjusted value), and the size representing the number of genes detected in each pathway. **(B-C)** Hallmark and WIKI-pathway (WP) analysis on sarcopenia patient cohort. NES, normalized enrichment score. **(D-E)** Heat map demonstrating the expression of genes associated with inflammation-related pathways and pathways related to muscle function across the young, aged and sarcopenia groups based on RNA-seq data. **(F)** Heat map demonstrating the correlation of inflammation-related pathways and pathways related to muscle function with the skeletal muscle microenvironment in the young, aged and sarcopenia groups based on RNA-seq data. Correlation coefficient values are colour-coded from blue (-0.4) to orange (0.4) using the colour bar on the right.

### Immune Cell Infiltration Features of sarcopenia

The enrichment of inflammatory signaling pathways is often accompanied by the infiltration of inflammatory immune cell populations. To further understand the microenvironment in sarcopenia, we categorized samples based on the distribution of macrophages, the dominant immune cell group in muscle tissue, which allowed us to observe the muscle immune microenvironment when macrophages were abundant or scarce. In the subsequent analysis, the samples were divided into two groups based on LYVE1 expression levels: LYVE1-high (above the median) and LYVE1-low (at or below the median). To investigate the immune microenvironment in different states, we calculated the percentages of immune cell types in each sample, and compared the results between the young, aged and sarcopenia groups as well as between the LYVE1-high and LYVE1-low groups (**Figure 6A**). Overall, there were no significant differences observed between the LYVE1-high and LYVE1-low group in most immune cell types. However, there were notable differences in B cells memory, NK cells resting and M2 macrophages, indicating activation of NK cells and B cells. Additionally, there was an increasing trend in monocytes in the LYVE1-high group (**Figure 6B**). To examine the variations in immune microenvironment compositions between the LYVE1-high and LYVE1-low groups, we utilized the ssGSEA algorithm to assess the enrichment score of genes associated with M0, M1 or M2-like macrophages as well as inflammation, and the expression of distinct immune cell types (**Figure 6C**). Our findings revealed that the majority of aged and sarcopenic individuals belonged to the LYVE1-high group, with an increase in inflammation-related gene expression. Additionally, the LYVE1-high group exhibited an increase in the expression of marker genes for M0, M1, and M2-like macrophages. This suggests that LYVE1 is a marker gene for tissue-resident macrophages in different stages of differentiation, given its specific expression in muscle macrophages (**Figure S1**). In the LYVE1-high group, we observed a higher presence of immune cells, FAP cells and endothelial cells in the skeletal muscle microenvironment. The correlations between cells in the LYVE1-high and LYVE1-low groups were found to be different. For instance, we found a negative correlation between M1 macrophages and NK cells in the LYVE1-low group, while a positive correlation was observed in the LYVE1-high group. Additionally, the correlation between type I and type IIa cells was stronger in the LYVE1-high group compared to the LYVE1-low group (**Figure 6D**). Stromal and basal scores showed a positive correlation with the expression of LYVE1+ macrophage, indicating an increase in basal cells and extracellular matrix in the presence of macrophages. Moreover, we investigated the correlation between LYVE1+ macrophages and FBN1+FAP cells as well as myeloid cells (**Figure 6E**). To validate our findings, we conducted histological staining (**Figure 6F**). PAS staining was utilized to visualize the distribution of glycogen in muscle tissue. The results clearly indicated that elderly individuals and patients with sarcopenia exhibited lower levels of muscle glycogen compared to young people, which aligns with previous studies(Consitt, Dudley, & Saxena, 2019). CD68 antibodies were employed to label macrophages. IHC analysis revealed a small number of macrophages scattered within the muscle interstitium of young samples. In contrast, a larger number of macrophages were found to be distributed in both aged and sarcopenic samples. These findings strongly suggest a correlation between macrophages and muscle fibrosis, with bone marrow-derived monocytes potentially serving as an important source of LYVE1+ macrophages.

**Figure 6.**
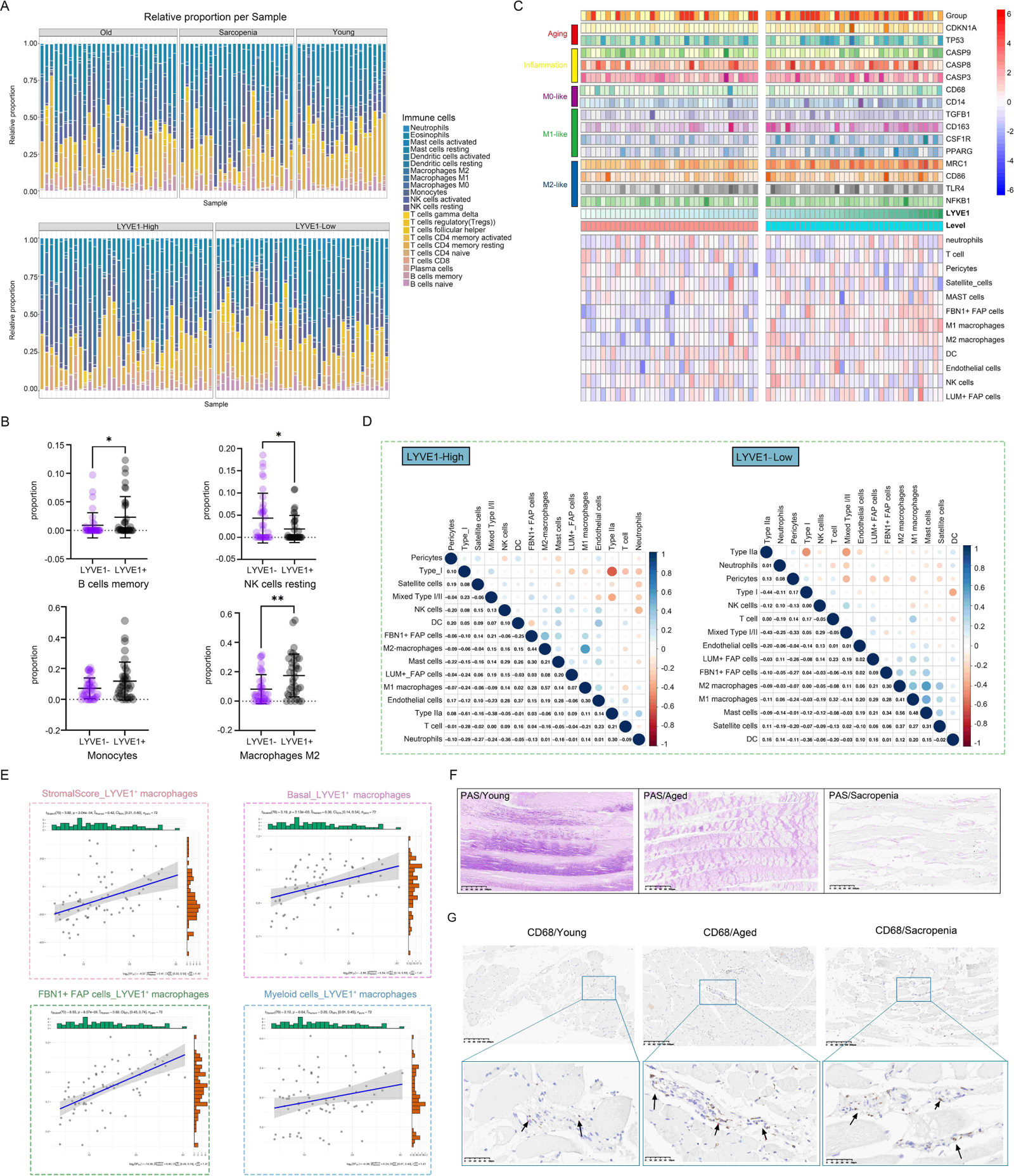
LYVE1 expression is strongly associated with the skeletal muscle immune microenvironment. **(A)** The proportion of tissue-infiltrating innate and adaptive immune cells assessed using the CIBERSORT algorithm is demonstrated in a stacked bar plot. The groups are categorized as young, aged, and sarcopenia patients, and further grouped based on high and low LYVE1 expression. **(B)** Cell proportion analysis of B cells memory, NK cells resting, monocytes and macrophages M2 between the LYVE1-high (LYVE1+) and LYVE1-low (LYVE1-) group (*p<0.05; **p< 0.01). **(C)** Heat map demonstrating the distribution of immune cell infiltration in the LYVE1-high and LYVE1-low groups generated based on the results, obtained from the ssGSEA and ESTIMATE algorithms and unsupervised hierarchical clustering. **(D)** Heat map showing the relationship between the infiltration levels of various innate and adaptive immune cells in the LYVE1-high and LYVE1-low groups. The Pearson correlation coefficients are provided, with blue indicating a positive correlation and orange indicating a negative correlation. **(E)** Scatter plot depicts the correlation of LYVE1+ macrophages with scores and cell expression in the skeletal muscle microenvironment. **(F)** Representative Periodic acid-Schiff (PAS) staining pathological images of tissue samples collected from the groups classified as young, aged, and sarcopenia (scale bar = 100 μm). **(G)** Representative images of immunohistochemical (IHC) staining for CD68 in human skeletal muscle tissue from the young, aged, and sarcopenia groups (scale bar = 100 μm). The black arrow points to the macrophages.

### Gene functional annotation analysis of modules associated with macrophages

Next, we conducted WGCNA to identify functional modules that were closely associated with macrophages. A total of 14 modules were detected, with the magenta and blue modules exhibiting the highest negative correlation with macrophages. Additionally, we selected two modules, pink and red, which showed a strong positive correlation with macrophages, for further investigation (**Figures 7A and B**). The correlation analysis revealed that the magenta and blue modules had a negative correlation with sarcopenia, while the pink and red modules showed a positive correlation with sarcopenia, indicating consistency between sarcopenia and macrophages. The pink module consisted of 299 genes primarily involved in cellular regulation, signaling pathways, intracellular mechanisms, and structural regulation. On the other hand, the red module was enriched in pathways associated with the extracellular matrix and fibrosis. Further analysis of genes in these two modules demonstrated significant enrichment in various pathways, including muscle structure development, DNA damage response, and cell cycle. The results were visualized using Cytoscape (**Figure 7C**). Next, we conducted an analysis on the modules that displayed a negative correlation with macrophages. Through Metascape analysis, we found that genes within the blue module were primarily enriched in pathways related to mitochondrial metabolism and function, and genes within the magenta module were found to be involved in various metabolic pathways and signal transduction pathways. Furthermore, when we merged these two modules and performed enrichment analysis, we observed that genes exhibiting a negative correlation with macrophages were significantly enriched in pathways related to mitochondria (**Figure 7D**).

**Figure 7.**
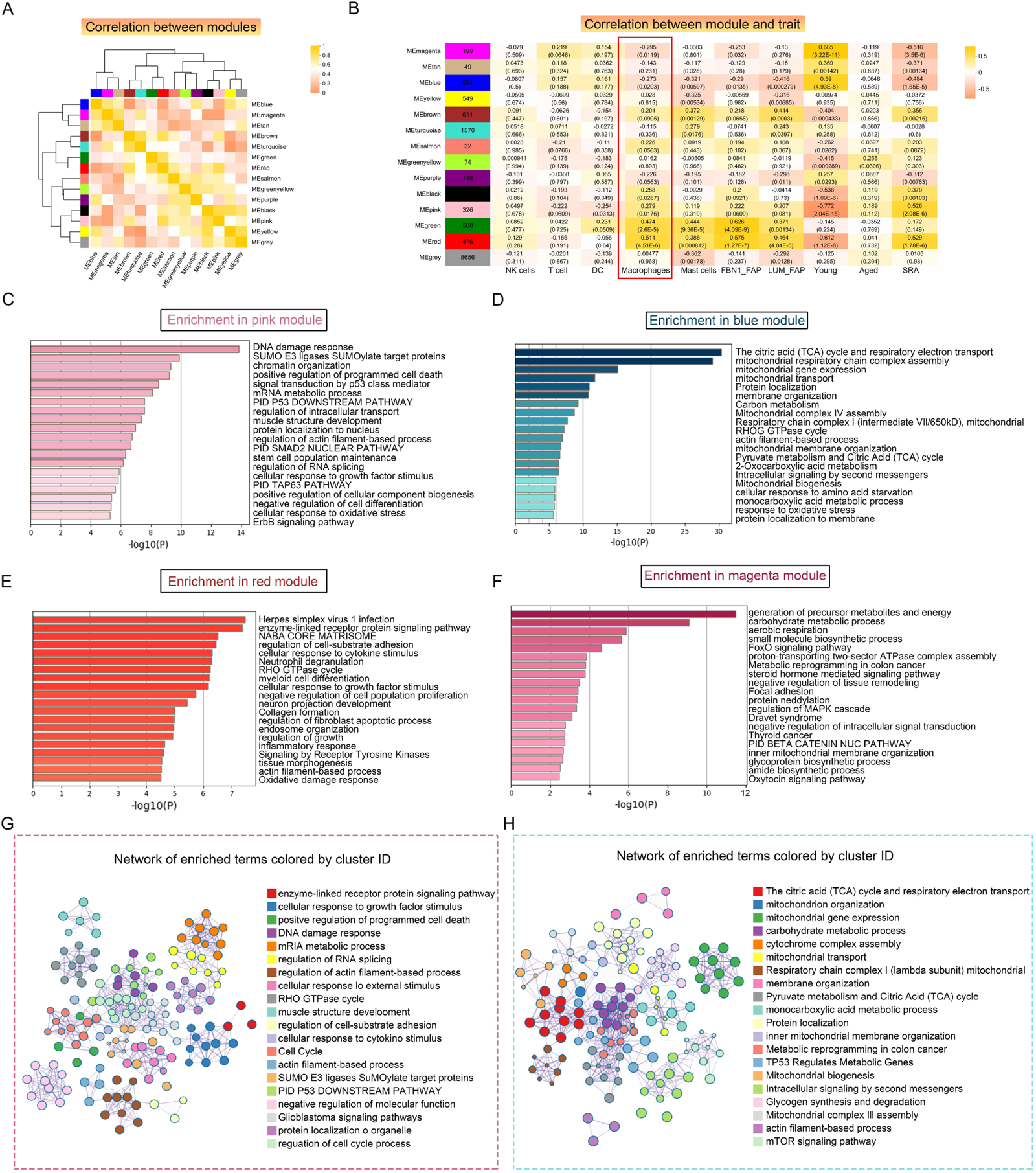
Weighted correlation network analysis (WGCNA) revealed unique functional gene modules associated with LYVE1+ macrophages. **(A)** Heat map demonstrating the Pearson correlation coefficients of eigengenes of co-expressed gene modules. A total of 14 modules were identified with highly correlated gene expression patterns. **(B)** Correlations between WGCNA modules and microenvironment cell type and age and disease state. Each cell represents Person’s correlation coefficient and p-value. **(C-F)** The results of Metascape for genes in the pink **(C)**, blue **(D),** red **(E)** and magenta **(F)** modules were represented using a horizontal bar chart. **(G-H)** Enrichment clustering network analysis in the Metascape database of red and pink merged modules **(G)** and blue and magenta merged modules **(H)**.

### Functional association network analysis of vital immune cells

We utilized GeneMINIA to predict the protein-protein interaction network between vital immune cells, particularly macrophages, in the skeletal muscle (**Figures 8A-B**). Next, we performed GO and KEGG analyses to interrogate the biological processes and pathways these relative genes involved in (**Figures 8C-D**). The identified immune cell-related pathways encompassed various immune system processes, signal transduction, adaptive immune response, as well as several T-cell differentiation and activation pathways. Additionally, the GO terms and KEGG pathways related to macrophages included pathways associated with cell signaling and tissue development, such as angiogenesis.

**Figure 8.**
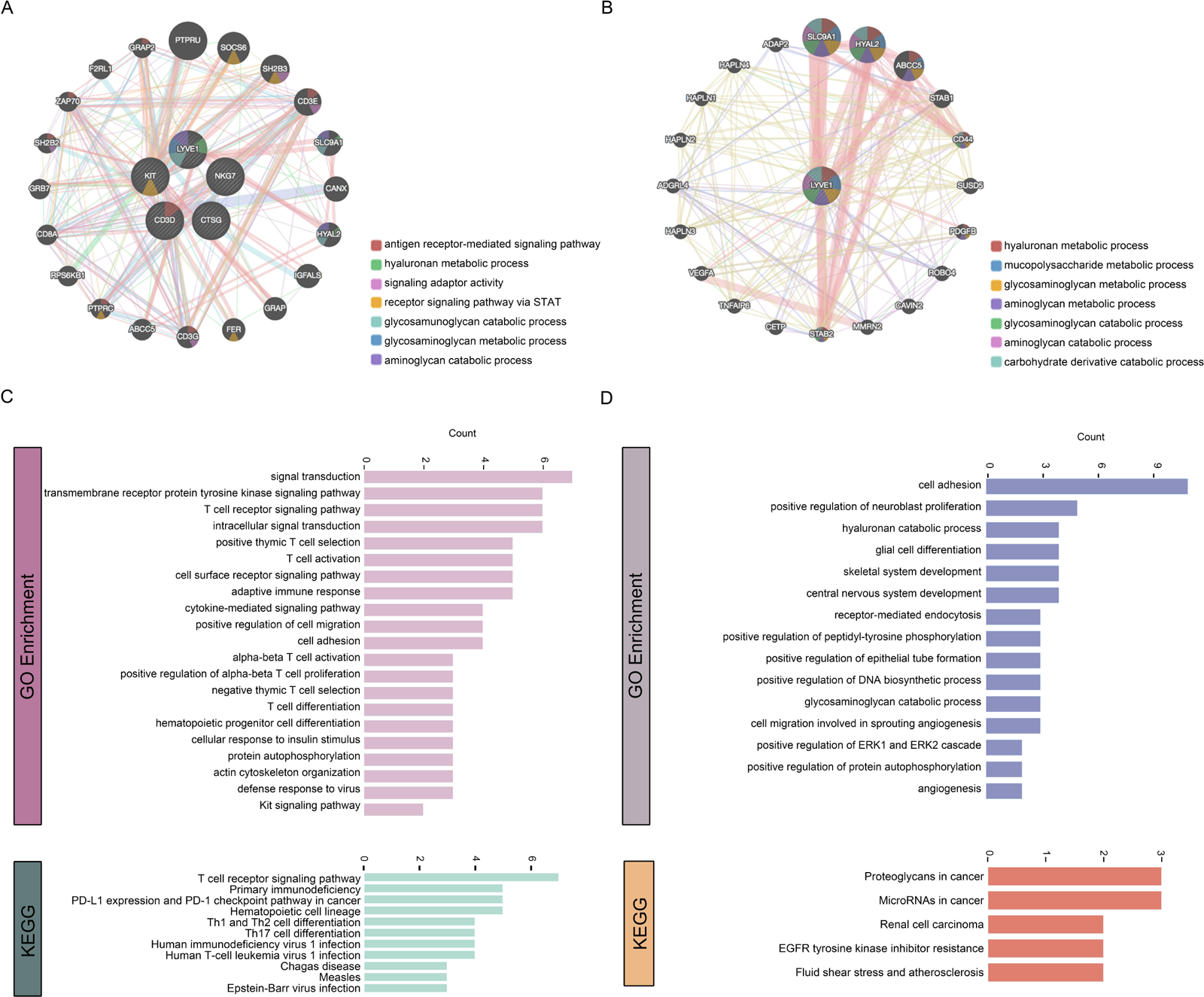
Functional association network analysis of vital immune cells. **(A-B)** Network inference carried out by examining the 5 genes that characterize T cells, NK cells, DCs, mast cells, and macrophages through GeneMANIA **(A)**. Additionally, only the LYVE1 gene considered **(B)**. The edges in the network representing interactions and color-coded based on the specific function of interaction occurring between the genes. Nodes represent genes associated with the hub genes. **(C-D)** GO and KEGG analyses on the associated genes to determine their functions.

## Discussion

Age-related decline in skeletal muscle mass and function is a multifactorial phenomenon, characterized by the loss of muscle fibers and the atrophy of remaining fibers(Ballak, Degens, de Haan, & Jaspers, 2014; Rezuş et al., 2020). Previous studies have highlighted the role of immune function changes in aging muscle(Nelke, Dziewas, Minnerup, Meuth, & Ruck, 2019; Reidy et al., 2019; Saini et al., 2016), but the specific alterations in the immune microenvironment of skeletal muscle with age have yet to be fully understood. In this study, we utilized single nuclei gene expression analysis in combination with bulk RNA-seq to investigate the differences in cell composition and immune microenvironment between young and aged skeletal muscle tissue. Since macrophages were found to be the predominant immune cell population, we further identified a specific marker gene LYVE1 for macrophages and clustered them into subgroups. Additionally, we explored changes in cell-cell interactions between aged and young muscle. Furthermore, by analyzing bulk RNA-seq data, we examined the gene expression signature, functional differences, and infiltration characteristics in the young, aged, and sarcopenia groups, as well as between the LYVE1-high and LYVE1-low groups. Our findings offer valuable insights into potential therapeutic strategies for sarcopenia, including targeting specific immune cell populations or modulating their activity. Notably, our results suggest a close association between macrophages and metabolic and functional alterations, as well as injury repair, in aging muscle.

Our analysis revealed that the sarcopenia group exhibited upregulation of several immune-related pathways, such as JAK-STAT, ERBB, and IL-2/4 signaling pathways, in comparison to young controls. Previous studies have also linked these pro-inflammatory pathways to age-related muscle atrophy(Abdelrahman et al., 2023; Ji et al., 2022). We also discovered a crosstalk between immune cells and various non-immune cells within the muscle microenvironment. The results indicated that immune cells received signals from fibroblasts, endothelial cells and other cell types, underscoring the significance of immune cells in the skeletal muscle microenvironment. Macrophages were found to be the predominant immune cell type in damaged muscles and played a crucial role in both the inflammation process and its resolution, contributing significantly to muscle repair(Chazaud, 2020; Siles, Ninfali, Cortés, Darling, & Postigo, 2019). Single-nucleus analysis revealed that the skeletal muscle immune microenvironment consisted of a diverse range of innate and adaptive immune cells, with macrophages comprising the largest proportion. Additionally, other immune cells were also involved in chronic muscle inflammation. Our research has revealed a significant correlation between DCs and macrophages. Similar to macrophages, DCs also function as potent antigen-presenting cells. When confronted with injury, DCs are activated to induce adaptive immunity, while macrophages initiate an inflammatory response(Linton & Thoman, 2014). Abnormal accumulation of DC has been reported in inflammatory myopathic muscles(Tournadre & Miossec, 2009). Within the muscle microenvironment, various immune cells exhibit distinct metabolic pathways and biological functions. M1 macrophages, for instance, are associated with the synthesis of collagen fibers and play a role in the TGF-beta signaling pathway. DCs are primarily involved in cell killing and sphingolipid metabolism signaling pathways. NK cells are predominantly enriched in the cytolysis pathway, while T cells are closely associated with skeletal muscle differentiation. Treg cells have been reported to enhance repair in chronically injured muscle because the depletion of Treg promotes the macrophages to switch from a pro-inflammatory to an anti-inflammatory phenotype(Kuswanto et al., 2016). Rebalancing the interactions between immune cells and other tissue cells, as well as among immune cells, may offer potential benefits for patients with sarcopenia.

As the most abundant type of immune cell in skeletal muscle, macrophages play a crucial role in maintaining muscle homeostasis. Resident macrophages in various tissues have unique chromatin landscapes and transcriptional profiles, which depend on their microenvironment and functional condition(Lazarov, Juarez-Carreño, Cox, & Geissmann, 2023). For this reason, there is currently no standardized macrophage signature. In the past, CD68 and F4/80 were established markers for resting M0 macrophages. Macrophage activation is triggered by extracellular and intracellular stimuli, leading to two main phenotypes: inflammatory (M1 and M2b) and wound resolution (M2a, M2c, and M2d)(Anders et al., 2022). These phenotypes exhibit distinct functional and metabolic characteristics. M1 macrophages are characterized by high expression levels of CD40 and CD86 and low expression levels of CD163, while functional phenotype of M2 macrophages are characterized by low expression levels of CD86 and high expression levels of CD163. Since macrophages in different tissues have varying functions and secrete different factors, this study identified LYVE1 as a marker for specific expression in macrophages within the skeletal muscle microenvironment. Similar findings were also reported by Krasniewski LK et al. in mouse muscles(Krasniewski et al., 2022). In this study, we performed bulk RNA-seq analysis and utilized LYVE1 as a macrophage marker. We divided the samples into high and low expression groups for LYVE1, and observed that the LYVE1+ macrophage enrichment group exhibited elevated expression levels of the classic M0, M1 and M2 subpopulation. Further transcriptomic analysis showed that the high-expression population of LYVE1+ macrophages was enriched in genes related to fibrosis and muscle development, suggesting the importance of macrophages in repairing damaged muscle.

Our study has also identified four distinct macrophage subpopulations based on marker gene expression, which may potentially serve different functional roles in the muscle immune environment and have diverse origins. The LYVE1^lo^MHC-II^h^i subpopulation exhibits enrichment in pathways associated with innate immune responses and antigen presentation, indicating a stronger pro-inflammatory phenotype. On the other hand, the LYVE1^hi^MHC-II^lo^ subpopulation demonstrates upregulation of metabolic pathways, suggesting its involvement in tissue remodeling. The LYVE1^lo^MHC-II^lo^TSPO+ subpopulation, which is enriched in extracellular matrix tissue genes, has been found to be closely associated with muscle fibrosis, and may play a role in the pathological accumulation of fibrosis observed in sarcopenia. Among these subpopulations, we noticed an increase in the expression of LYVE1^hi^MHC-II^lo^ subpopulation in aged muscle. In a study conducted on mice, it was discovered that the LYVE1^lo^MHC-II^hi^ cells originated entirely from hematopoietic stem cells (HSCs), while the LYVE1^hi^MHC-II^lo^ subpopulation consisted of an equal proportion of cells derived from HSCs and non-HSCs(X. Wang et al., 2020), which may suggest their different origins.

Currently, targeting immune cell-related pathways within the microenvironment has emerged as a crucial therapeutic strategy for sarcopenia. For instance, supplementing MANF protein can promote the transformation of macrophages from a pro-inflammatory phenotype to a repair phenotype, thus facilitating aging muscle regeneration and regulating inflammation and tissue homeostasis(Sousa et al., 2023). IL-15 has also been identified as a promising therapeutic target for mitigating inflammation-induced skeletal muscle atrophy(O’Leary, Wallace, Bennett, Tsintzas, & Jones, 2017). Further understanding the distinct functions and origins of macrophage subpopulations could offer valuable insights for developing treatments that target specific pro-inflammatory or fibrotic phenotypes.

This study has a few limitations. Firstly, the sample size could be increased to provide more robust results. Additionally, further experiments are needed to verify the efficiency of LYVE1 as a marker gene for skeletal muscle macrophages. While macrophages in the skeletal muscle microenvironment have been categorized, the specific subpopulations that are closely associated with the development of sarcopenia and their corresponding targets still require further investigation.

Overall, in this study, we have identified LYVE1 as a marker gene for tissue-resident macrophages in the skeletal muscle microenvironment across different ages and disease states. Our findings suggest a correlation between LYVE1+ macrophage expression and sarcopenia, as well as significant functional and metabolic differences between the high and low expression groups. Furthermore, our snRNA-seq analysis has uncovered new clustering patterns in human skeletal muscle macrophages. Additionally, we have observed variations in the skeletal muscle microenvironment among aged and young individuals and those with sarcopenia. Our research provides valuable insights into potential strategies for treating sarcopenia by targeting macrophages.

## Methods

### Data preparation

Single-nuclei RNA transcriptome, bulk RNA transcriptome data and corresponding clinical information were obtained from the Gene Expression Omnibus (GEO) database (https://www.ncbi.nlm.nih.gov/geo/). The dataset GSE167186 contained 72 Bulk RNA-seq samples from young, aged and sarcopenic subjects, as well as 17 single-nuclei RNA-seq samples from young and aged subjects, with a total of 143,051 nuclei included. LIMMA package was used to normalize the RNA-sequencingdata(Ritchie et al., 2015).

### Single-nuclei RNA-seq data analysis

The quality control and downstream analysis of sn-RNA-seq data were performed using the Seurat R package (http://satijalab.org/seurat/). Low-quality nuclei were filtered using the cutoffs nFeature_RNA > 500 & nFeature_RNA < 4000 & percent.mt < 18 & nCount_RNA > 500 & nCount_RNA < 10000, resulting in101,862 cells remaining. Thet-SNE algorithm was utilized for cell cluster generation(Becht et al., 2018). Cell types within the clusters were manually annotated based on cell markers from original publications and well-known markers using SingleR(Aran et al., 2019). Bubble charts and violin charts were employed to visualize the expression of marker genes in each cluster.

We screened pathways related to functions, metabolism, and immunity processes in skeletal muscle using the kyoto encyclopedia of genes and genomes (KEGG) dataset (https://www.kegg.jp/kegg/) and the Gene Ontology (GO) dataset (https://biit.cs.ut.ee/gprofiler/). Gene set variation analysis (GSVA) was performed using the GSVA R package to characterize the differentially expressed pathways of each cluster(Hänzelmann, Castelo, & Guinney, 2013).The average expression values of genes for each cell type were used as input data. Cell-cell communications (CCCs) among cell clusters in the skeletal muscle microenvironment, especially among immune cell clusters were identified and visualized using the CellChat package. The enriched ligand-receptor (LR) interactions between a macrophage subpopulation and other clusters were detected using the Cellchat package based on known LR pairs.

### Bulk RNA-seq data analysis

The Gene Set Enrichment Analysis (GSEA) (https://www.gsea-msigdb.org/gsea/) and pathway signal score calculation was conducted using KEGG, Hallmark, and WIKI-pathway gene sets. Differentially expressed pathways with a normalized (NOM) P-value of <0.05 were kept for further functional analysis. To create heat maps based on gene expression in young and aged skeletal muscle tissues, we utilized the ggplot2 R package (http://ggplot2.tidyverse.org/) with fragments per kilobase of transcript per million sequenced reads (FPKM) data. The single-sample gene set enrichment analysis (ssGSEA) algorithm was then employed to evaluate the enrichment score of each pathway(Barbie et al., 2009). We used the pheatmap R package to generate the heatmap for correlation coefficients of mitochondria-related and muscle-function related pathways, and muscle microenvironment genes. For the correlation analysis, we used the Spearman correlation test

The ESTIMATE package was used to calculate ESTIMATE, immune, and stromal scores for the RNA-seq data of young, aged, and sarcopenia samples(Yoshihara et al., 2013), and the enrichment scores calculated by the ssGSEA analysis were used to evaluate immune-related gene activity and the relative abundance of infiltrating immune cells. We then performed deconvolution on our RNA-seq dataset using the CIBERSORT algorithm (http://cibersort.stanford.edu) to estimate the relative fraction of immune cells. The correlation analysis among different immune cell types was conducted using the ggstatsplot package, and the results were visualized using the ggplot2 package.

To identify groups of highly correlated genes, we used the unsupervised method called weighted gene correlation network analysis (WGCNA)(Langfelder & Horvath, 2008). The eigengenes from each module were used to assess the relationship between the modules and immune cell types, as well as clinical information. For functional annotation analyses, we utilized the online tool Metascape (http://metascape.org)(Zhou et al., 2019). The network of enriched term subsets was visualized using Cytoscape v3.10.1 (https://cytoscape.org/). GeneMINIA (http://www.genemania.org) was used to predict the functional association network of hub genes(Warde-Farley et al., 2010).

### Tissue samples collection and cohort description

Adult human skeletal muscle tissue samples for immunostaining were obtained from 18 patients. These patients were categorized into three groups: young patients (aged 35 years or younger), aged patients without sarcopenia (aged 60 years or older), and aged patients with sarcopenia (aged 60 years or older). Demographic information, including age, gender, medical history, and pathological diagnosis, was collected for each patient. All individual participants have provided the informed consents and the study was approved by the Ethics Committee of Ruijin Hospital, Shanghai Jiao Tong University School of Medicine and registered with the Chinese Clinical Trial Registry.

### Histological analysis

Fresh muscle samples were fixed in 10% neutral buffered formalin overnight at room temperature. The samples were then embedded in paraffin. The paraffin-embedded tissue sections, 4μm thick, were air-dried and further dried at 75°C for 2 hours and then dewaxed. Hematoxylin and eosin (H&E) staining was carried out using a CoverStainer (Dako, Germany) following standard procedures. Immunohistochemical (IHC) procedures were performed on an automated Leica Bond RX staining platform (Leica Biosystems, Welzlar, Germany) following the selected protocol for the BOND Polymer Refine Detection kit (DS9800, Leica Biosystems).

For a list of primary antibodies, please refer to Supplemental table S1. For Periodic acid-Schiff (PAS) staining, the sections were incubated in Periodate solution for 5 minutes, and then stained with Schiff reagent using a PAS Staining Kit following a protocol suggested by the manufacturer. Images were captured using a Nikon Eclipse Ni-U microscope equipped with a Nikon DS-Ri digital camera. Immunofluorescence (IF) staining was performed on skeletal muscle tissue samples as previously described(Vetter, Nicolau, Bradley, Frair, & Flanigan, 2022). The primary antibodies used, along with their dilutions, are listed in Supplemental table S2. Cell nuclei were labeled with DAPI (blue). Representative fluorescence images were captured on a Leica TCS SP8 MP confocal microscope (Leica Microsystems, Wetzlar Germany).

### Statistical analysis

All data were expressed as the mean ± standard deviation (SD). R (version 4.2.3) and GraphPad Prism (version 8) were used for statistical analysis. The t-test or one-way ANOVA test was used for comparison between groups was performed with Statistical correlation between variables was evaluated by the Spearman correlation test. P-values less than 0.05 were considered statistically significant.

## Acknowledgenments

Not applicable.

## Additional information Funding

We received no financial support for the research, authorship, and publication of this article.

## Author contributions

Linhui Shen, Conceptualization, Methodology, Data curation, Writing – original draft; Yuan Zong, Methodology, Data curation, Software, Writing – original draft; Jiawen Zhao, Data curation, Software, Visualization, Writing – original draft; Yi Yang; Methodology, Data curation, Software, Writing – original draft; Lei Li, Investigation, Visualization; Ning Li, Investigation, Visualization; Yiming Gao, Validation; Xianfei Xie, Supervision; Qiyuan Bao, Conceptualization, Methodology, Writing – review & editing; Liting Jiang, Conceptualization, Methodology, Writing – review & editing; Weiguo Hu, Conceptualization, Methodology, Writing – review & editing.

## Ethics

All procedures involving patients in our study were approved by the Ethics Committee of Ruijin Hospital, Shanghai Jiao Tong University School of Medicine.

## Additional files

### Data availability

All data are available in the main text or Supplementary file 1. The previously published dataset that support the findings of this study is GEO: GSE167186

The following previously published dataset was used:

| Author(s) | Year | Dataset title | Dataset URL | Database and Identifier |
| --- | --- | --- | --- | --- |
| Perez K, Melov S, Ciotlos S | 2022 | Single-nuclei profiling of aged skeletal muscle | https://www.ncbi.nlm.nih.gov/geo/, accession number GSE167186 | NCBI Gene Expression Omnibus, GSE167186 |

