## Supplemental Table1 for "Characterizing the skeletal muscle immune microenvironment for sarcopenia using transcriptome analysis and histological validation"

Table S1 Primary antibodies detailed information

| antigen | dilution/concentration | Cat. # | | Company and Nation |
| --- | --- | --- | --- | --- |
| CD45 | 1:100 | 60287-1-Ig | Proteintech, China | |
| CD3 | undiluted | GA503 | Dako, Denmark | |
| CD68 | undiluted | GA613 | Dako, Denmark | |
| CD56 | 1:100 | IR628 | Dako, Denmark | |
| CD117 | undiluted | KIT-0029 | MXB Biotechnologies, China | |
| α-SMA | 1:500 | GB12044 | Servicebio, China | |
